# Association between allostatic load and accelerated white matter brain aging: findings from the UK Biobank

**DOI:** 10.1101/2024.01.26.24301793

**Authors:** Li Feng, Zhenyao Ye, Zewen Du, Yezhi Pan, Travis Canida, Hongjie Ke, Song Liu, Shuo Chen, L. Elliot Hong, Peter Kochunov, Jie Chen, David K.Y. Lei, Edmond Shenassa, Tianzhou Ma

**Author notes:** Correspondence: Tianzhou Ma, Department of Epidemiology and Biostatistics University of Maryland School of Public Health, Edmond Shenassa, Department of Epidemiology and Biostatistics University of Maryland School of Public Health.

## Abstract

White matter (WM) brain age, a neuroimaging-derived biomarker indicating WM microstructural changes, helps predict dementia and neurodegenerative disorder risks. The cumulative effect of chronic stress on WM brain aging remains unknown. In this study, we assessed cumulative stress using a multi-system composite allostatic load (AL) index based on inflammatory, anthropometric, respiratory, lipidemia, and glucose metabolism measures, and investigated its association with WM brain age gap (BAG), computed from diffusion tensor imaging data using a machine learning model, among 22 951 European ancestries aged 40 to 69 (51.40% women) from UK Biobank. Linear regression, Mendelian randomization, along with inverse probability weighting and doubly robust methods, were used to evaluate the impact of AL on WM BAG adjusting for age, sex, socioeconomic, and lifestyle behaviors. We found increasing one AL score unit significantly increased WM BAG by 0.29 years in association analysis and by 0.33 years in Mendelian analysis. The age- and sex-stratified analysis showed consistent results among participants 45-54 and 55-64 years old, with no significant sex difference. This study demonstrated that higher chronic stress was significantly associated with accelerated brain aging, highlighting the importance of stress management in reducing dementia and neurodegenerative disease risks.

## Introduction

Human brain aging is characterized by continuous microstructure transformations, including a reduction in cerebral integrity of grey and white matter compartments and cognitive decline(1, 2). Specifically, white matter microstructural integrity can be an early biomarker for the prodromal stage of dementia and other neurodegenerative disorders(3–5). We propose that allostasis, a dynamic physiological adaptation to chronic exposure to stressors(6), provides a working model to understand how chronic physical and psychosocial stressors may affect brain aging. In our investigation, we employed a machine learning algorithm(7–9), leveraging multiple fractional anisotropy tract measurements obtained from diffusion tensor imaging data to predict white matter (WM) Brain Age Gap (BAG), representing the difference between an individual’s chronologic and biological brain age. The mechanisms by which chronic stress impacts brain aging are not fully understood. Hence, we examined the association between chronic stress, quantified by Allostatic Load (AL), and biological brain aging, assessed through WM BAG using multiple statistical methods, from linear regression to potential causality inferring Mendelian Randomization, inverse probability weighting, and doubly robust methods.

Allostatic load was developed to quantify the “wear and tear” on the body, stemming from the continuous activation of stress responses via the hypothalamic–pituitary–adrenal axis and the sympathetic-adrenal-medullary system(10). A cross-sectional study of Lothian Birth Cohort (b. 1936) showed evidence of a positive association between AL and brain age by using T1-weighted MRI scans of gray matter and white matter among cognitively healthy older adults (mean age 73 years) residing in Edinburgh Scotland(11). Other investigations showed a similar positive association between elevated AL and white matter volume and integrity with limited sample size from 349 to 731 participants in mainly local cross-sectional cohorts(12–15) (See Table S1 for details). The selection of a brain aging indicator is essential. Recent studies highlight the efficacy of advanced machine learning techniques applied to neuroimaging data, offering the age-adjusted BAG estimates with minimal predictive biases(16, 17) proven to be linked to physical and brain health (9, 18–22). The single scalar metric BAG is a robust and easily interpretable outcome, simplifying the challenges of handling multiple variables and addressing dependencies typically encountered in complex multivariate brain imaging models(7, 8, 16).

To meet the challenge of establishing a link between AL and WM BAG while accounting for the multifactorial nature of WM BAG influenced by genetic, environmental, biological, and psychosocial factors(23–25), besides a linear regression model, we employed a Mendelian randomization method. Mendelian randomization method has been increasingly applied in observational studies by leveraging genetic variants as instrumental variables to establish a potential causal relationship between exposure and outcome(26). Compared to the previous methods, genetic variants are considered useful instrumental variables because they are less prone to be influenced by reverse causation or confounding factors, making them particularly strong for causal inference(27). Additionally, we considered a dichotomized AL risk and applied inverse probability weighting(28–30) and doubly robust methods to estimate the effect of AL on BAG for more robust inference(31).

In this study, we evaluated the association between AL and WM BAG among the general population as well as stratified by sex and age in the UK Biobank cohort. We hypothesized that an increase in chronic stress burden would accelerate white matter brain aging. The estimated effects derived from our study shed light on the long-term physiological toll of chronic stress on brain health. This research is a critical step for developing early interventions that can reduce the global burden of dementia by addressing chronic stress risk factors before they lead to irreversible brain damage.

## Methods

### Study population and variables

The data used in the present study were from the UK Biobank, a large-scale population-based longitudinal cohort study recruiting over 500 000 individuals aged 40 to 69 years in 22 recruitment centers across the UK at baseline from 2006-2010 and collecting comprehensive physical, environmental, genomic, and health data. Beginning in 2014, the brain imaging phenotypic data (N=∼40k) were collected(32). Imaging visit (instance 2, 2014+) where our outcome of interest WM BAG was measured happened six years after the baseline where our exposure the AL biomarkers were measured, providing the basis for evaluating the potentially causal relationship.

In this study, we focus on the non-pregnant, predominantly European white (self-reported) population in UK Biobank. We study white participants only to avoid bias of imbalanced data and reduce cross-population heterogeneity in training the brain age prediction model and we have shown in our previous studies that the white-only model estimates brain age well with high prediction accuracy (9, 21) The participants who had all five AL biomarkers at baseline (N = 348 361) were included to calculate the AL index. The participants who had fractional anisotropy data available but did not have extreme WM hyperintensities (N=30 375) were included to estimate the age-adjusted WM BAG in the testing dataset. Extreme WM hyperintensities can significantly alter brain structure and skew the brain age prediction(33). After merging, the final analytic sample includes N=22 951 participants for the analysis using linear regression, inverse probability weighting and doubly robust methods. For one-sample Mendelian Randomization (MR) analysis, we further merged the final analytical sample with genetic data and excluded participants with kinship, excessive heterozygosity, and sex discordance based on genetic data, resulting in an analytical sample of N=15 678 participants. These exclusion criteria are standard in Genome-Wide Association Studies (GWAS) to improve data reliability, maintain the integrity and accuracy of the genetic analysis(34, 35), and ensure more robust MR analyses. The details of the inclusion and exclusion criteria, data processing and analytical steps can be found in Figure 1.

**Figure 1.**
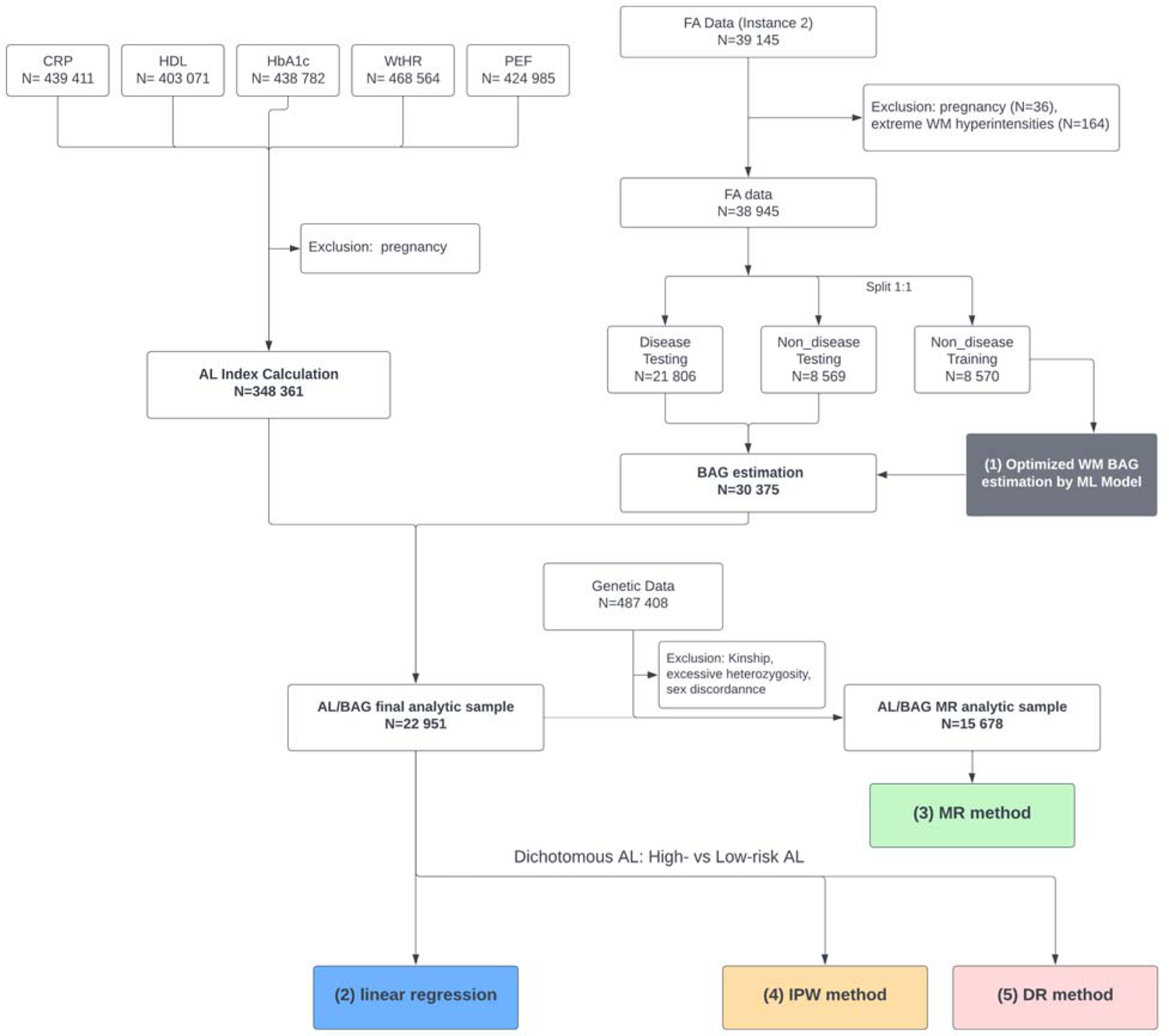
Flow chart of participants selection and data processing procedures. Flowchart of our main analysis procedures and the number of subjects included at each step of the analysis. The main analyses are in three parts: (1) Estimation of age bias corrected white matter (WM) brain age gap (BAG) using machine learning model (Gray); (2) Linear association analysis between allostatic load (AL) index and BAG (Blue); (3) mendelian randomization (MR) method (green) to evaluate the potencial causal effect of AL on BAG; (4) inverse probability weighting (IPW) method (orange); (5) doubly robust (DR) method (pink).

### Allostatic load (AL)

Our AL index is comprised of five biomarkers measured at baseline representing distinct physical systems: C-reactive protein (mg/L) representing immunological/inflammatory, waist-to-height-ratio representing anthropometric, peak expiratory flow (litres/min) representing respiratory, high-density lipoprotein cholesterol (mmol/L) representing lipidemia, glycated hemoglobin (mmol/mol) representing glucose metabolism. These five biomarkers were found to be available in most studies and predict mortality and longer, more elaborate AL batteries in recent meta-analysis(36).

Individuals at the highest quartile for C-reactive protein, waist-to-height-ratio, and glycated hemoglobin and the lowest quartile for high-density lipoprotein cholesterol and peak expiratory flow were marked as 1 and indicated as health risk among each 5-year and sex-specific distribution and otherwise as 0. Utilizing a combination of five distinct biomarkers representing various physiological systems, our AL index provides a comprehensive assessment of the cumulative effects of chronic stress on an individual’s health, offering a precise understanding of AL’s impact on brain aging(37). We further validated our proposed AL index by 1) testing its correlation with the subclinical health outcomes/measures (depressive score, self-reported health, left/right-hand grip strength, usual walking pace, and duration of walking for pleasure) as positive control and 2) testing its correlation with non-health-related variables (water intake each day, milk type used, usual side of head for mobile phone use) as negative control. Results of the additional validation tests can be found in Supplementary Table S2 and S3.

### White matter brain age gap (WM BAG)

Our main outcome, the WM BAG, was computed based on the fractional anisotropy measures collected from diffusion tensor imaging data from the UK Biobank imaging assessment (instance 2, 2014+)(9). This study focuses on 39 WM fractional anisotropy tracks that cover multiple brain regions (see Table S4). Following the ENIGMA protocol, the average value for each white matter tract in the brain was computed using Track-Based Spatial Statistics analysis applied to the diffusion tensor imaging fractional anisotropy images(38). The fractional anisotropy measurement, which falls within the range of 0 to 1, reflects the level of anisotropy in a diffusion process and the integrity of the cortical white matter(39).

We applied a machine learning model (random forest regression) to predict brain age and estimate BAG from fractional anisotropy data. We followed a similar data splitting scheme in previous studies to split the data into a training set (N=8570) and a testing (N=30 375) (9, 40). The training set includes healthy participants only (excluding who were previous or current smokers, had hypertension, CVD, diabetes, or brain diseases, etc.). We employed random forest regression with fractional anisotropy features as predictors and chronological age as the outcome. The model’s parameters were optimized using a 5-fold cross-validation, focusing on maximizing the correlation between chronological age and estimated brain age, and minimizing mean absolute error (MAE). This approach also identified key fractional anisotropy features influencing brain aging. The optimized model was then applied to the testing set for brain age prediction. We calculated the WM BAG by subtracting the chronological age from the predicted brain age. To address age-dependent bias noted in previous studies(22, 41), we used linear regression for bias correction, assessing the correction method’s performance via MAE(42). The adjusted WM BAG from the testing set was the primary outcome in subsequent analyses.

### Potential confounders and instrumental variables (IVs)

We included potential confounding factors from baseline in the UK Biobank, including sociodemographic factors: age, sex (females and males), education (A levels/AS levels or equivalent, College or University degree, CSEs or equivalent, NVQ or HND or HNC or equivalent, O levels/GCSEs or equivalent, and other professional qualifications), Townsend Deprivation Index (an area-based score of social deprivation (accounting for unemployment, overcrowding, non-car ownership, and non-home ownership)), household income (less than 18k, 18k-31k, 31k-52k, 52k-100k, more than 100k); lifestyle/behavior factors (e.g. diet, smoking, physical activity; all were calculated based on American Heart Association Life’s Essential 8’s definition, ranged 0-100 for each score (Higher score means healthier behavior))(43). We assumed missing at random for these covariates and performed multiple imputations by chained equations method to impute the covariates data(44).

For MR analysis, we further incorporated the genotype data to select the genetic variants as candidate instrumental variables (IVs). The genotype data were assayed with UK BiLEVE Axiom Array and with UK Biobank Axiom Array(45, 46). We performed quality control of genotype data and only kept the genetic variants with: minor allele frequency ≥ 0.01, Hardy-Weinberg equilibrium exact test *P* value ≥ 0.001, missing genotype rate ≤ 0.05 and missingness per individual ≤ 0.2. In the Mendelian randomization analysis, we further filtered out subjects who had missing genotypes for more than two valid SNPs and used random imputation to impute the missing genotypes(47).

## Statistical analysis

### Descriptive statistics

Variables of baseline characteristics are shown as % (N) if categorical, mean (SD) if continuous. The mean (SD) distribution of the AL index and WM BAG by baseline categorical variables was compared using an ANOVA test. Pearson correlation tests were applied to test the association between the continuous variables and the AL index and WM BAG. *P* and r values were shown for the significance of both tests and the Pearson correlation coefficient, respectively (Table 1). The baseline characteristics for the age- and sex-stratified groups are detailed in Table S5. Table S6 compares the baseline characteristics of participants included in the AL/BAG final analytic sample vs. those excluded due to pregnancy or extreme WMH; Table S7 compares the baseline characteristics of those included in the MR analytic sample vs. excluded due to kinship, excessive heterozygosity, and sex discordance.

**Table 1.**
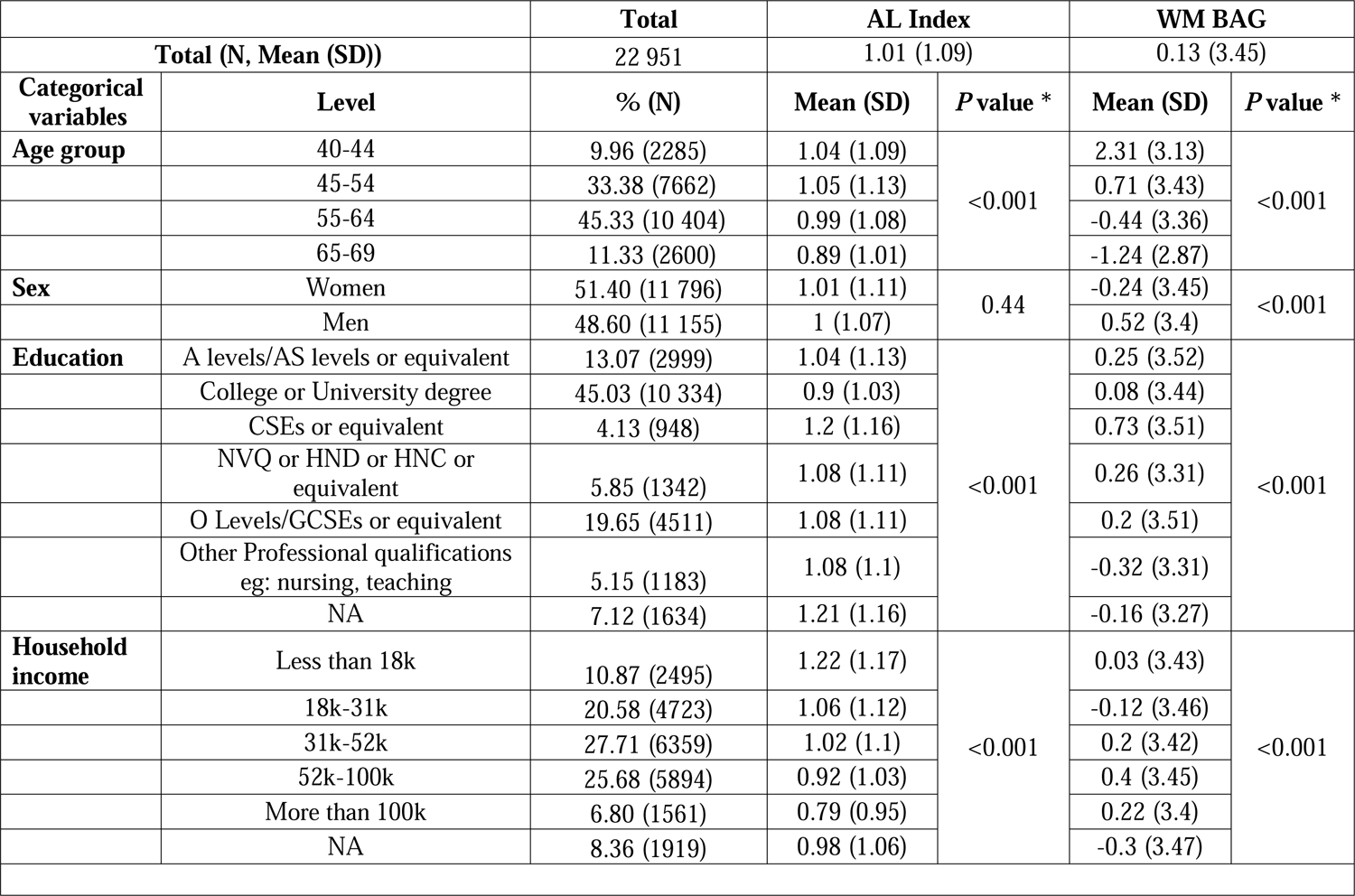
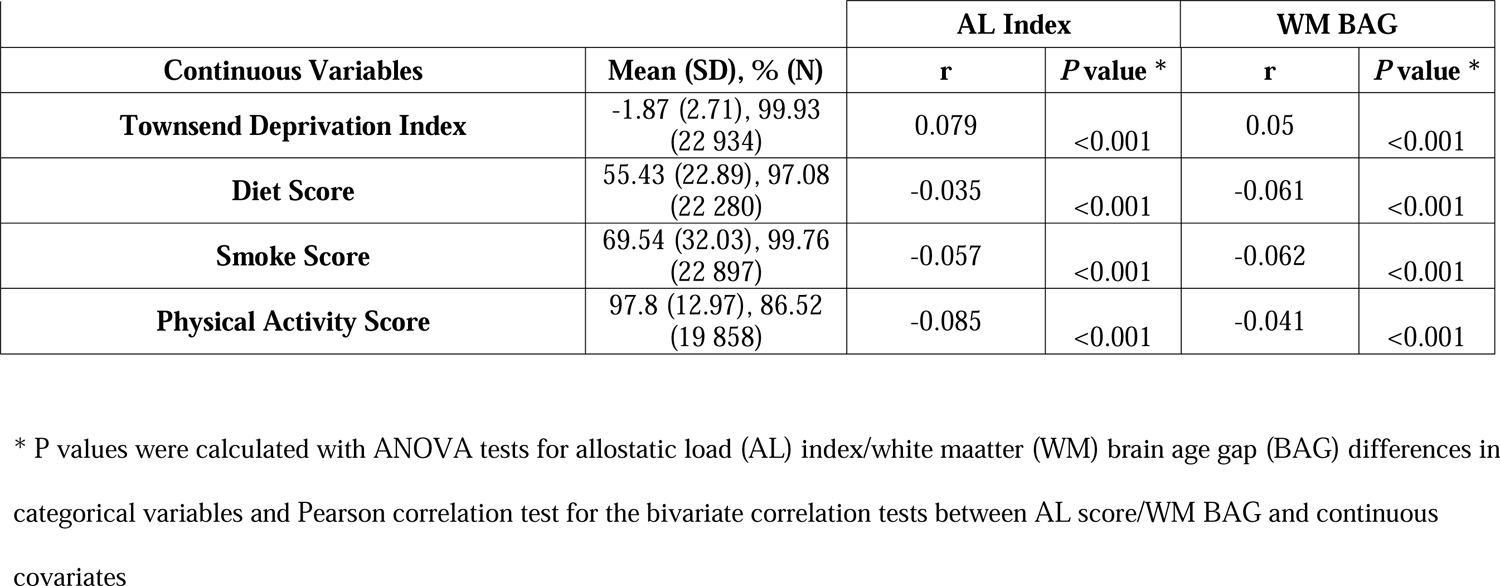
Characteristics of Participants for linear regression, inverse probability weighting, and doubly robust estimation.

|  |  | **Total** | **AL Index** |  | **WM BAG** |  |
| --- | --- | --- | --- | --- | --- | --- |
| **Total (N, Mean (SD))** |  | 22 951 | 1.01 (1.09) |  | 0.13 (3.45) |  |
| **Categorical variables** | **Level** | **% (N)** | **Mean (SD)** | **P value \*** | **Mean (SD)** | **P value \*** |
| **Age group** | 40-44 | 9.96 (2285) | 1.04 (1.09) | <0.001 | 2.31 (3.13) | <0.001 |
|  | 45-54 | 33.38 (7662) | 1.05 (1.13) |  | 0.71 (3.43) |  |
|  | 55-64 | 45.33 (10 404) | 0.99 (1.08) |  | -0.44 (3.36) |  |
|  | 65-69 | 11.33 (2600) | 0.89 (1.01) |  | -1.24 (2.87) |  |
| **Sex** | Women | 51.40 (11 796) | 1.01 (1.11) | 0.44 | -0.24 (3.45) | <0.001 |
|  | Men | 48.60 (11 155) | 1 (1.07) |  | 0.52 (3.4) |  |
| **Education** | A levels/AS levels or equivalent | 13.07 (2999) | 1.04 (1.13) | <0.001 | 0.25 (3.52) | <0.001 |
|  | College or University degree | 45.03 (10 334) | 0.9 (1.03) |  | 0.08 (3.44) |  |
|  | CSEs or equivalent | 4.13 (948) | 1.2 (1.16) |  | 0.73 (3.51) |  |
|  | NVQ or HND or HNC or equivalent | 5.85 (1342) | 1.08 (1.11) |  | 0.26 (3.31) |  |
|  | O Levels/GCSEs or equivalent | 19.65 (4511) | 1.08 (1.11) |  | 0.2 (3.51) |  |
|  | Other Professional qualifications<br>eg: nursing, teaching | 5.15 (1183) | 1.08 (1.1) |  | -0.32 (3.31) |  |
|  | NA | 7.12 (1634) | 1.21 (1.16) |  | -0.16 (3.27) |  |
| **Household income** | Less than 18k | 10.87 (2495) | 1.22 (1.17) | <0.001 | 0.03 (3.43) | <0.001 |
|  | 18k-31k | 20.58 (4723) | 1.06 (1.12) |  | -0.12 (3.46) |  |
|  | 31k-52k | 27.71 (6359) | 1.02 (1.1) |  | 0.2 (3.42) |  |
|  | 52k-100k | 25.68 (5894) | 0.92 (1.03) |  | 0.4 (3.45) |  |
|  | More than 100k | 6.80 (1561) | 0.79 (0.95) |  | 0.22 (3.4) |  |
|  | NA | 8.36 (1919) | 0.98 (1.06) |  | -0.3 (3.47) |  |

|  |  | **AL Index** |  | **WM BAG** |  |
| --- | --- | --- | --- | --- | --- |
| **Continuous Variables** | **Mean (SD), % (N)** | **r** | **P value \*** | **r** | **P value \*** |
| **Townsend Deprivation Index** | -1.87 (2.71), 99.93<br>(22 934) | 0.079 | <0.001 | 0.05 | <0.001 |
| **Diet Score** | 55.43 (22.89), 97.08<br>(22 280) | -0.035 | <0.001 | -0.061 | <0.001 |
| **Smoke Score** | 69.54 (32.03), 99.76<br>(22 897) | -0.057 | <0.001 | -0.062 | <0.001 |
| **Physical Activity Score** | 97.8 (12.97), 86.52<br>(19 858) | -0.085 | <0.001 | -0.041 | <0.001 |

\* P values were calculated with ANOVA tests for allostatic load (AL) index/white matter (WM) brain age gap (BAG) differences in categorical variables and Pearson correlation test for the bivariate correlation tests between AL score/WM BAG and continuous covariates

### Association analysis between AL index and BAG

We used four statistical models to investigate the association between AL and WM BAG (Figure 2) among the general population as well as stratified by sex and age groups (early middle-aged (40–44), late middle-aged I (45–54), late middle-aged II (55–64), and early older adults (65–69)) in the UK Biobank cohort. We first applied a linear regression model to test the association between WM BAG and AL, controlling for the aforementioned confounders. We also tested for age and sex interactions with AL in both the linear regression and one-sample MR to determine whether these factors significantly influenced the association between AL and WM BAG.

**Figure 2.**
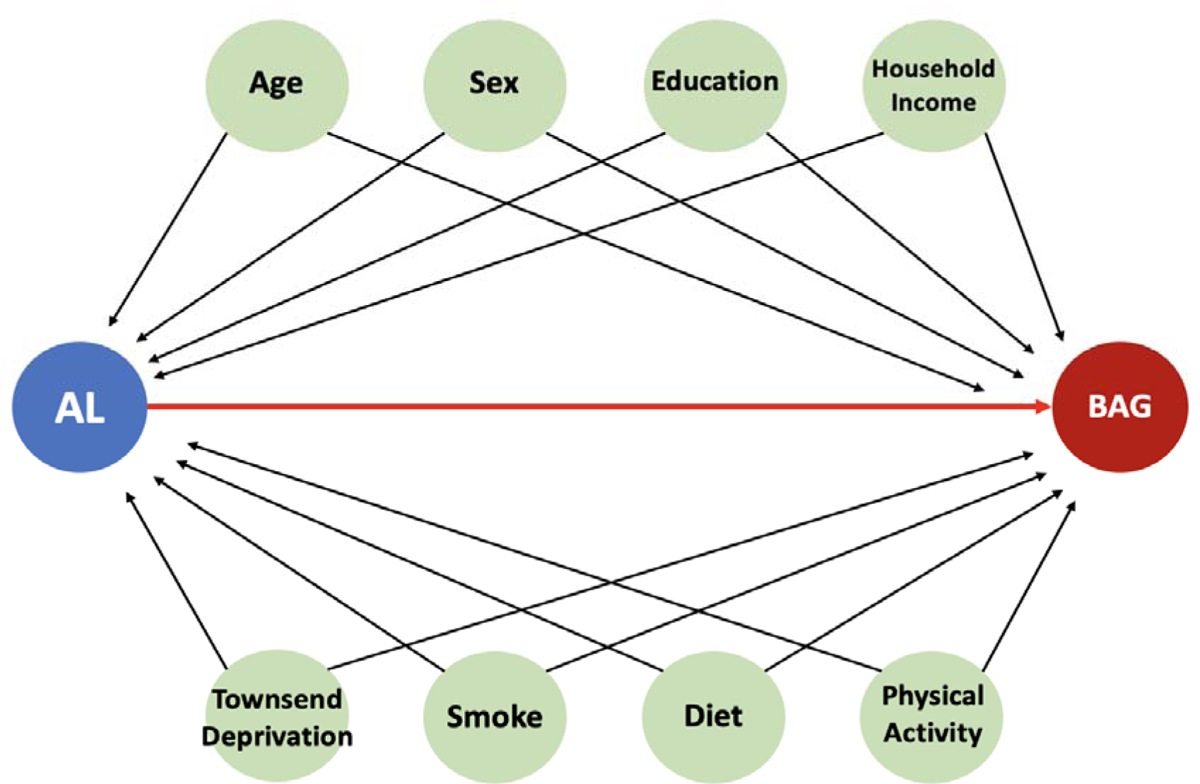
Directed Acyclic Graphs to investigate the potential causal relationship between allostatic load (AL) and white matter (WM) brain age gap (BAG). Multiple models: model 1 (Multiple linear regression), model 2 (one-sample Mendelian Randomization), model 3 (inverse probability weighting method), model 4 (doubly robust method). Confounders: age, sex, education, household income, Townsend deprivation score, smoke score, diet score, and physical activity score.

Association analysis using linear regression models does not directly infer causality. We then performed a one-sample MR analysis by treating genetic variants as IVs to evaluate the potential causal relationship between AL and WM BAG following the guidelines by Burgess et al. (2023)(48). First, from a larger non-overlapping sample (N=217 606) with valid AL index but no fractional anisotropy data in the UK Biobank, we performed a GWAS (35) (*P* < 5X 10^-8^) adjusting for population stratification followed by linkage disequilibrium clumping (r^2^=0.5 within window 250 kb) using Plink (version 1.9, www.cog-genomics.org/plink/1.9/) to select the variants associated with AL index as candidate IVs (IV assumption (i))(49). In one-sample mendelian randomization analysis, we further removed variants significantly associated with the covariates (false discovery rate adjusted *P* >0.05; IV assumption (ii)) and then removed the horizontal pleiotropic variants by regressing BAG on AL index with the genetic variables (false discovery rate adjusted *P* >0.1; IV assumption (iii)). We annotated the final selected IVs using ANNOVAR (https://annovar.openbioinformatics.org/en/latest/) and used R packages “*gwascat*” to find relevant traits associated with these IVs. We also validated the IV selection by testing the IV effect on AL using the first stage F-statistics and the IV x age or IV x sex interaction effects. We mainly used the inverse-variance weighted (IVW) MR method and other popular MR methods (simple median, weighted median, penalized weighted median, inverse variance weighted, Egger, etc.) were also applied for sensitivity analysis.

To further verify our findings, we split the participants into AL high-risk (AL index ≥2; N=6208) and low-risk group (AL index <2; N=16 743) and estimated the effect of AL risk on BAG using the inverse probability weighting (IPW) and doubly robust (DR) estimation methods. We also conducted sensitivity analyses by removing the outlier participants (N=1223) with extreme propensity scores, or redefining the high-risk AL group (AL index ≥ 1, 3, 4, as representing the top 59.28%, 10.68%, 3.14% of the population, respectively) and rerunning IPW method for robustness.

All statistical analyses were conducted using R (version 4.0.5)(50). R packages, including “*Hmisc*” (version 5.1.0), “*MICE*” (version 3.14.0), “*ipw*” (version 1.2), “*MendelianRandomization*” (version 0.5.1)(51) were used to perform multiple imputation by chained equations, inverse probability weighting, doubly robust, and Mendelian randomization analyses. Except for IV selection, *P*<0.05 was regarded as statistically significant for all other analyses unless otherwise specified. For IV selection in MR analyses, Benjamini-Hochberg method was used to adjust for multiple comparisons(52).

## Results

The optimal random forest regression model selected 16 fractional anisotropy measures to predict brain age (Table S2). The age-adjusted predicted brain age achieved a Pearson correlation coefficient (R)= 0.89, mean absolute error (MAE) = 2.74 years in the testing dataset (Figure S1), implying an excellent prediction performance of the proposed machine learning model. In the final analytical sample of 22 951 participants (mean age 55.37, 51.40% women), the mean AL index (ranging from 0-5) is 1.01 (SD, 1.09), and the mean of WM BAG is 0.13 (SD, 3.45).

Participants with a lower AL index were older, had higher education, more household income, lower Townsend deprivation index, and higher diet, smoke, and physical activity scores, but no sex difference (Table 1). We also observed that older groups, especially older females, had lower WM BAG, AL index, Townsend deprivation index, and higher diet, smoke, and physical activity scores (Table 1 and Table S5). The baseline characteristics are largely alike between the included and the excluded participants (Table S6-7) and no clear selection bias has been observed in our study.

Overall, all the four models showed significant positive relationship between AL and WM BAG (Table 2). In the linear regression model, when adjusting for age, sex, education, household income, Townsend Deprivation Index, smoking, diet, and physical activity, WM BAG is estimated to increase by 0.29 years (106 days, 95% CI: 0.25 to 0.33 years, *P*<0.001) per one AL score increment.

**Table 2.** Results of 1) association analysis by linear regression model between continuous allostatic load (AL) index and white matter (WM) brain age gap (BAG), 2) one-sample Mendelian randomization (MR) analysis between continuous AL index and WM BAG, 3) inverse probability weighting (IPW) between High/low-risk AL groups and WM BAG, 4) doubly robust (DR) between High/low-risk AL groups and WM BAG among overall participants. All models were adjusted for age, sex, education, Townsend deprivation index, household income, smoke score, diet score, and physical score.

| Level | Model | Exposure | Estimate | Lower 95%<br>CI | Upper 95%<br>CI | *P* value |
| --- | --- | --- | --- | --- | --- | --- |
| Overall | Linear Regression | AL index (Continuous) | 0.29 | 0.25 | 0.33 | <0.001 *** |
|  | One-sample MR | AL index (Continuous) | 0.33 | 0.01 | 0.65 | 0.042 * |
|  | IPW | High-risk AL (Binary) | 0.60 | 0.51 | 0.69 | <0.001 *** |
|  | DR | High-risk AL (Binary) | 0.60 | 0.50 | 0.70 | <0.001 *** |

P-value significance:
\* p<0.05 \*\* p<0.01 \*\*\* p<0.001

In one-sample MR model, the GWAS analysis identified 2636 genetic variants significantly (*P*< 5 X10^-8^) associated with AL index (see Manhattan plot in Figure S2). A final set of 238 variants were identified as valid IVs after checking the IV assumptions (Table S8). All these IVs had first stage F-statistics above the acceptable threshold and showed no significant interactions with age or sex (Table S9). Many of these IVs were found to be associated with biomarkers including C-reactive protein, hemoglobin and high-density lipoprotein cholesterol collected in GWAS Catalog(53), aligning well with the five biomarkers used to define our multi-system AL index (Table S9). Using these IVs, we found an overall significant positive effect of AL index (0.33 years/120 days, 95% CI=0.01 to 0.655 years per one AL score increment, *P*=0.042) on WM BAG (Table 2).

For dichotomized AL risk group comparison, both the IPW and DR methods showed that participants with high-risk AL were older in WM BAG than those with low-risk AL (IPW: 0.60 years/219 days, 95% CI: 0.51 to 0.69 years, *P* <0.001; DR: 0.60 years/219 days, 95% CI: 0.50 to 0.70 years, *P* <0.001), confirming a consistent finding of the positive relationship between AL and WM BAG (Table 2).

The results of the age- and sex-stratified analyses can be found in Figure 3, Table S10. No significant interactions between AL and age or sex were observed (Table S11). In the age-stratified analysis, the positive association between AL index and WM BAG was consistent in all four age strata (early middle age, late middle age I, late middle age II, late adulthood) from linear regression and inverse probability weighting method but not one-sample Mendelian randomization method, while we observed that the late middle-aged participants (45-54 years, 55-64 years) had the most significant effects. Sex showed no significant difference overall, but some sex disparities were found in different age strata.

**Figure 3.**
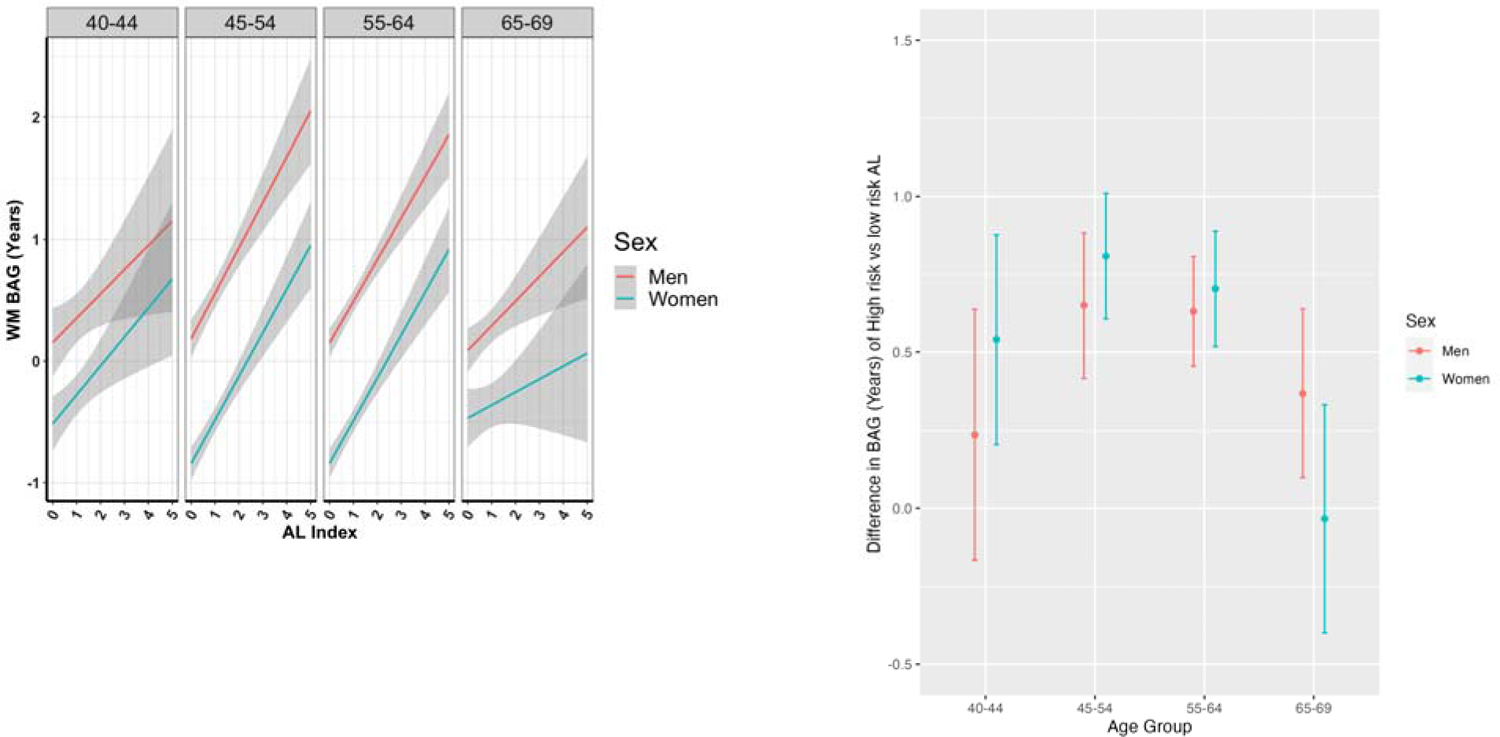
(Left) Associations between adjusted white matter (WM) BAG (brain age gap) and allostatic load (AL) index (continuous) by sex and age group (Gray area, 95% CI). (Right) inverse probability weighting (IPW)-handled association analyses between WM BAG and High-risk AL (binary) by sex and age group. The estimate represents the absolute WM BAG difference between High-risk AL and Low-risk AL.

The sensitivity analysis results from other MR methods matched well with that by the main IVW method (Table S12). The results were not materially changed when removing extreme propensity scores (Figure S3, Table S13), or redefining high-risk vs. low-risk AL groups (Table S14) for IPW method.

## Discussion

This study reveals a consistently positive association and potentially causal relationship between AL and WM BAG among the participants of White ethnicity aged 40-69 in the UK Biobank across a range of statistical models, with the strongest effect observed among participants aged 45-64 years old. To the best of our knowledge, this study marks the first exploration of the potential causal relationship between AL and brain aging in a large community-dwelling population, encompassing both the overall population and age- and sex-specific groups. Our proposed AL index represents a universal and consensus set of biomarkers from multiple systems that capture the physiological ‘wear and tear’ and were further validated by using positive and negative controls. It is noteworthy that our machine learning-based BAG estimation exhibits remarkable predictive accuracy. These results illuminate the positive relationship between chronic stress and brain aging, offering insights into potential strategies for promoting healthy brain aging and mitigating dementia and neurodegenerative diseases burden.

Previous studies(12–15, 54, 55) have identified a negative association between cortisol levels (i.e., serum, hair) and brain volume, changes in the microstructure of gray or white matter, and impaired cognitive functions. For chronic stress, our results are consistent with evidence indicating a positive link between chronic stress (AL index) and WM volume(12, 13, 54), WM tracks(15, 55), and WM microstructure(14). In contrast to earlier studies that used multiple singular measurements, WM BAG offers a holistic estimation of brain aging that is derived from 39 brain-wide white matter fractional anisotropy tracks using a machine learning model, providing a single measure of brain aging that is predictive of cognitive decline(23) and standardized for use across diverse studies(56).

The complex mechanism underlying the impact of chronic stress on brain aging and the microstructure of white matter remains an active area of research. At the core of this relationship is the dysregulation of the hypothalamic-pituitary-adrenal axis, a primary characteristic of an impaired adaptive stress response commonly observed in brain aging(57). Chronic stress precipitates a cascade of endocrine disruptions, causing sustained elevation of cortisol—a glucocorticoid linked to deleterious effects on cerebral architecture and functionality, manifesting as compromised white matter integrity(58, 59). This may be partly due to disrupted myelination process of oligodendrogenesis that maintained repair white matter. Chronic stress targets the prefrontal cortex, amygdala, and hippocampus(60), and can alter oligodendrogenesis and thus the balance between myelin damage and repair(61, 62). Structural changes in white matter can compromise its fundamental role in facilitating signal transmission between brain regions(63), hence indicating the changes in white matter may be a mediator between chronic stress and cognitive function and emotion regulation. The relation between chronic stress and persistent systemic inflammation, alongside an increase in oxidative stress, is potentially an additional pathway contributing to the brain’s vascular alterations, neurodegeneration(64), and accelerated aging of the white matter.

Collectively, these findings suggest that chronic stress acts through both hormonal dysregulation and inflammatory pathways may elicit changes that are detrimental to the brain’s white matter, potentially hastening the natural aging process. This study has provided empirical evidence that AL contributes to WM BAG. However, the precise mechanisms by which AL influences the biological brain age remains to be further elucidated, which is crucial for developing targeted interventions.

This study exhibits several strengths in design and analysis compared to other assessments of the impact of AL on brain aging. Firstly, we leveraged a large-scale UK Biobank cohort and conducted various statistical analyses, enabling us to unravel a potential causal relationship between chronic stress and brain aging across overall and different age and sex groups. Secondly, informed from the meta-analysis(36), we incorporated five robust indicators into one AL index, representing vital physiological systems validated from positive and negative controls (Supplementary Tables S3 and S4). Thirdly, we employed an optimal machine learning model to calculate the WM BAG and adjusted age bias, resulting in an accurate estimation of biological brain age. Fourthly, we utilized multiple robust causality inference methods, including Mendelian randomization, inverse probability weighting and doubly robust approaches, which mitigated the impact of confounding variables and reverse causation, thus enhancing the validity of the study. Lastly, we conducted multiple sensitivity analyses on AL measurements and High-risk AL definitions, as well as alternative Mendelian randomization analyses, ensuring the reliability and validity of our findings.

The study has certain limitations that offer opportunities for future research. First, the UK Biobank comprises a relatively healthy population, which may skew the sample towards healthier individuals and potentially introduce selection bias(65). The general population may include a larger proportion of subjects more vulnerable to the damaging effects of AL (e.g. those with mental illness(37)). This could compromise the generalizability of our results and potentially lead to an underestimation of the association between allostatic load and brain aging. Second, the construct of AL index in this study was limited to biomarkers available through the UK Biobank. To date, over 50 clinical and non-clinical biomarkers were found to define AL index before(36), yet no consensus has been reached regarding the optimal set of biomarkers. Future research could leverage high-throughput omics technologies to explore the potentially common molecular underpinnings and mechanism of chronic stress in the body, mitigating the controversy surrounding AL definition. Lastly, by design of UKB cohort, we cannot further track each participant for their change in chronic stress and brain aging over time, ongoing longitudinal cohorts such as Adolescent Brain Cognitive Development study and Baltimore Longitudinal Study of Aging are to be investigated in the future study to justify the causal relationship(66, 67).

## Conclusion

In conclusion, in this pioneering examination of the association and potential causal relationship between chronic stress and brain aging using a robust set of statistical methods, we observed a positive relationship between allostatic load and WM BAG among the participants of White ethnicity aged 40-69 in the UK Biobank, which highlights the critical role of chronic stress management in preventing early-onset cognitive decline and advancing public health strategies against dementia and neurodegenerative diseases.

## Author Contributions

L.F. took the lead in performing the analysis and wrote the manuscript. Z.Y. and Z.D. performed the one-sample Mendelian randomization analysis. T.M. and E.S. developed the idea, supervised the project and took the lead in editing the manuscript. Y.P., T.C., H.K.,S.C., E.H., P.K., J.C., D.L. contributed to manuscript writing and polishing. All authors provided critical feedback and helped to shape the research, analysis, and manuscript.

## Data Availability Statement

The raw genetic and phenotypic data used for this study can be found in the UK Biobank (http://www.ukbiobank.ac.uk/)

